# A Comprehensive Study of GPT-4V’s Multimodal Capabilities in Medical Imaging

**DOI:** 10.1101/2023.11.03.23298067

**Authors:** Yingshu Li, Yunyi Liu, Zhanyu Wang, Xinyu Liang, Lingqiao Liu, Lei Wang, Leyang Cui, Zhaopeng Tu, Longyue Wang, Luping Zhou

**Author notes:** Equal Contribution (Yingshu Li, Yunyi Liu, Zhanyu Wang).

## Abstract

This paper presents a comprehensive evaluation of GPT-4V’s capabilities across diverse medical imaging tasks, including Radiology Report Generation, Medical Visual Question Answering (VQA), and Visual Grounding. While prior efforts have explored GPT-4V’s performance in medical imaging, to the best of our knowledge, our study represents the first quantitative evaluation on publicly available benchmarks. Our findings highlight GPT-4V’s potential in generating descriptive reports for chest X-ray images, particularly when guided by well-structured prompts. However, its performance on the MIMIC-CXR dataset benchmark reveals areas for improvement in certain evaluation metrics, such as CIDEr. In the domain of Medical VQA, GPT-4V demonstrates proficiency in distinguishing between question types but falls short of prevailing benchmarks in terms of accuracy. Furthermore, our analysis finds the limitations of conventional evaluation metrics like the BLEU score, advocating for the development of more semantically robust assessment methods. In the field of Visual Grounding, GPT-4V exhibits preliminary promise in recognizing bounding boxes, but its precision is lacking, especially in identifying specific medical organs and signs. Our evaluation underscores the significant potential of GPT-4V in the medical imaging domain, while also emphasizing the need for targeted refinements to fully unlock its capabilities.

## 1 Introduction

Large Language Models (LLMs) have consistently demonstrated remarkable prowess across various domains and tasks (Touvron et al., 2023; OpenAI, 2023; Anil et al., 2023). The ongoing pursuit of enhancing LLMs’ capacity for visual comprehension has spurred the emergence of a new research area: Large Multimodal Models (LMMs) (Ye et al., 2023; Li et al., 2023b; Awadalla et al., 2023). The basic approach has been to either fine-tune the visual encoder to align with a fixed pre-trained LLM or to use a vision-language model to convert visual input into textual descriptions that can be understood by the LLM. These applications are all based solely on the use of the LLM and do not really explore the visual capabilities of the LLM. GPT-4V, a cutting-edge Large Multimodal Model (LMM) incorporating visual understanding capabilities, is constructed as an evolution of state-of-the-art Large Language Models (LLMs). This model is trained on an extensive corpus of multimodal data. Yang et al. conducted a comprehensive case study to assess GPT-4V’s performance in general-purpose scenarios, revealing its robust visual comprehension ability (Yang et al., 2023b). Moreover, LMMs have been widely used in the medical field (Wang et al., 2023b; Singhal et al., 2023). The introduction of visual capabilities into GPT-4V opens up opportunities for an in-depth examination of its potential in the domain of medical multimodality. So this paper will evaluate the main image, and multi-modal tasks in the medical imaging field based on GPT-4V.

The main contribution of this paper is to explore the capabilities of GPT-4V on medical image analysis. We selected the 3 main medical multimodal tasks, **Radiology Report Generation, Medical Visual Question Answering**, and **Medical Visual Grounding**, to assess GPT-4V’s performance in the context of medical images. Our evaluation encompassed *standard benchmarks* and comparative analysis against current state-of-the-art models. Furthermore, we conducted in-depth case studies using representative examples for each task, enhancing our comprehension of GPT-4V’s capabilities in medical image understanding.

## 2 RElated Work

### 2.1 RADiology Report Generation

Radiology report generation has emerged as a prominent research area within the domain of medical image analysis in recent years. While similar to image captioning (Vinyals et al., 2015; Xu et al., 2015; Pan et al., 2020), this task presents heightened complexity due to the extended length of medical reports and the increased difficulty in identifying medical anomalies within images, due to data imbalance issues. Numerous research has relied on encoder-decoder architectures to address this task. The research can be grouped into two primary research directions. The first direction concentrates on enhancing the model’s architecture to facilitate improved extraction of visual features and the generation of high-quality medical reports. For example, Li et al. used a hierarchical architecture to generate reports with normality and abnormality respectively (Li et al., 2018). Similarly, Liu et al. employed a hierarchical structure to initially generate topics and subsequently produce related sentences (Liu et al., 2019). With the prevailing of the transformer (Vaswani et al., 2017), Chen et al. introduced a transformer-based model, enhancing it with relational memory and memory-driven conditional layer normalization to enhance image feature recognition and capture crucial report patterns (Chen et al., 2020). Another research direction is to solve the data bias problem by incorporating external knowledge information. Zhang et al. constructed a predefined medical knowledge graph to augment the model’s ability to capture valuable medical information (Zhang et al., 2020). To further enrich this supplementary knowledge, Li et al. developed a dynamic approach that enables real-time updates to the knowledge graph (Li et al., 2023c).

Furthermore, in recent times, there has been a surge in radiology report generation methods leveraging Large Language Models (LLMs). These approaches leverage the capabilities of large language models to generate long-text content and utilize abundant knowledge sources to enhance the quality of radiology reports. Wang et al. employs Llama2 to elevate the quality of the generated reports. To achieve effective image-text alignment, the image embeddings are mapped to the feature space of the Llama2 (Touvron et al., 2023) via a visual mapper to ensure uniform dimensionality (Wang et al., 2023b).

### 2.2 Visual Question Answering

Visual Question Answering (VQA) (Jiang et al., 2020; Wu et al., 2019) has solidified its stature as a paramount domain, striving to empower machines to decipher visual content and respond to pertinent natural language inquiries. Given a pair comprising an input image and a correlated question, the VQA model is engineered to generate the corresponding answer. A plethora of previous scholarly works have delved into VQA, revealing four critical components within these models: the image encoder, the text encoder, a fusion method, and either a generator or a classifier, contingent upon the model’s architectural design. The nature of the posed questions bifurcates into two categories based on the answer type: the close-end type (Nguyen et al., 2019; Finn et al., 2017; Eslami et al., 2021) and the open-end (Ambati & Dudyala, 2018; Khare et al., 2021) type. Predominantly, models address these two categories distinctly; they typically employ a classification-based approach for close-end types, whereas for open-end types, a generation-based method is utilized. Nevertheless, a select number of studies have attempted to integrate both question types within a singular model (Ren & Zhou, 2020). A notable example is the Q2ATransformer (Liu et al., 2023), which simultaneously tackles both varieties of questions, amalgamating the strengths of classification-based and generation-based methodologies, and subsequently achieving exemplary accuracy across both question types.

With the emergence of Large Language Models (LLMs), there has been a substantial influx of research leveraging LLMs to augment the linguistic inferencing capabilities of VQA (Li et al., 2023a). Moreover, certain studies have pioneered the use of LLMs for facilitating continuous questioning in VQA. The introduction of models such as GPT-3.5 has led to the generation of more LLM-based datasets, mitigating the issue of data scarcity (Pellegrini et al., 2023). The advent of GPT-4V marks a significant milestone, as it incorporates image comprehension capabilities directly into the LLM framework. This eliminates the need for VQA systems to translate all tasks into a language understandable by traditional LLMs. With the ability to process multimodal inputs seamlessly, the evolution of LLMs has opened new horizons for research and development in VQA. This paper endeavors to elucidate the application of GPT-4V in the realm of medical image-based VQA, exploring its potential and implications in this specialized field.

### 2.3 Visual Grounding

Visual grounding (Kamath et al., 2021) stands as a pivotal field at the crossroads of computer vision and natural language processing. Essentially, this task requires interpreting an image while taking into account a relevant textual description of an object, which could range from a single sentence or caption to a more elaborate description. The end goal is to produce a bounding box that accurately outlines the designated object. Given its critical role in integrating visual and textual information, visual grounding has established itself as a crucial application in the realm of multimodal interactions.

With the emergence of extensive language modeling, there has been a noteworthy blend of visual grounding techniques with Large Language Models (LLMs) (Peng et al., 2023; Zhao et al., 2023). In a conventional setup, data from bounding boxes, obtained through visual grounding, is fed into the LLM as a segment of the prompt. This approach steers the LLM towards making the right assessments. However, the debut of GPT-4V marks a significant transformation in this workflow. It eliminates the requirement for crafting prompts manually, allowing users to directly input images and text, and in turn, directly obtain the related bounding box outputs. This advancement simplifies the process, removing the need for extra steps and intermediaries.

The majority of visual grounding research primarily centers on general imagery, with scant attention paid to the realm of medical images. This disparity could stem from a noticeable dearth of datasets specifically tailored for medical visual grounding. The MS-CXR dataset, a recent published visual grounding dataset, makes some improvement of medical image visual grounding application, some publications (Huang et al., 2023; Sun et al., 2023a;b) comes out base on it. Nevertheless, even as this dataset becomes more widely recognized, there remains a limited body of academic work exploring its potential and applications, highlighting a crucial area for future research and development.

In this paper, we will embark on a comprehensive review of GPT-4V’s applications within the domain of medical visual grounding, exploring its capabilities, impact, and potential avenues for future research and development.

## 3 Radiology Report Generation

The exponential growth of radiological imaging data has imposed an escalating burden on radiologists, leading to a heightened risk of diagnostic errors with potentially severe consequences. Consequently, there is a growing demand for automated radiology report generation, which is anticipated to alleviate the workload of radiologists and mitigate diagnostic inaccuracies. The rapid advancements in artificial intelligence, particularly in the domains of computer vision and natural language processing, have made automated medical report generation a feasible reality (e.g., Chen et al., 2020; 2021; Liu et al., 2021; Wang et al., 2023a). A prominent challenge in automated medical report generation is long text generation. Presently, large language models (LLMs) (e.g., Touvron et al., 2023; Chowdhery et al., 2022) have gained widespread prominence and demonstrate a strong proficiency in generating long text. Furthermore, LLM-based large multi-modal models (LMMs) (e.g., Zhu et al., 2023; Wu et al., 2023) possess a notable capability for multi-modal content generation. While LMMs show potential in multi-modal content generation, their efficacy in specialized tasks like radiology report generation is yet to be fully explored. The accuracy and reliability of such reports are paramount, making it crucial to evaluate LMMs in this domain rigorously. In the following sections, we examined the GPT-4V’s capability for generating radiology reports using distinct prompt strategies and the dataset.

### 3.1 Evaluation

This section presents an evaluation of the GPT-4V model’s capacity for medical report generation. We employ the MIMIC-CXR dataset (Johnson et al., 2019) for assessment. The model is tasked with generating diagnostic reports for given medical images. To facilitate comparison with established methodologies(e.g., Chen et al., 2020; Yang et al., 2021a; Liu et al., 2021; Wang et al., 2022b; 2023a), we employ widely recognized metrics, specifically BLEU scores (Papineni et al., 2002), ROUGE-L (Lin, 2004), METEOR (Banerjee & Lavie, 2005), and CIDEr (Vedantam et al., 2015), to gauge the quality of the generated reports.

Our evaluation focuses on the model’s performance with the MIMIC-CXR testset. Each evaluation instance comprises a single medical image coupled with a carefully crafted text prompt as the input.

#### 3.1.1 Dataset: MIMIC-CXR

MIMIC-CXR, the largest publicly available dataset in this domain, includes both chest radiographs and unstructured textual reports. This dataset comprises a total of 377,110 chest X-ray images and 227,835 corresponding reports, obtained from 64,588 patients who underwent examinations at the Beth Israel Deaconess Medical Center between 2011 and 2016. To facilitate fair and consistent comparisons, we followed the official partitioning provided by MIMIC-CXR, resulting in a test set containing 3,858 samples.

#### 3.1.2 Prompt Design Strategies for Evaluation

To better activate the capabilities of the GPT-4V, we explored different prompt design strategies, including zero-shot and few-shot approaches. In the **zero-shot** scenario, we provided examples without reference reports, while in the few-shot way, we explored three different prompt settings: (1) two normal reports **(Few-shot normal examples prompt)**, (2) two abnormal reports **(Few-shot abnormal examples prompt)**, and (3) one normal report paired with one abnormal report **(Few-shot mixed examples prompt)**. Our extensive evaluation of these prompt strategies unveiled that the inclusion of both a normal and an abnormal example consistently led to the generation of higher-quality reports. Consequently, we employed prompts with one normal and one abnormal example **(Few-shot mixed examples prompt)** to evaluate GPT-4V on MIMIC-CXR benchmark.

#### 3.1.3 Overview of Prompt Methods

Our primary objective is to evaluate the baseline performance of GPT-4V in medical report generation. Consequently, we focused solely on the **zero-shot prompt** and the **few-shot prompt**, avoiding the use of complex techniques like chain-of-thought (Wei et al., 2022b) or ensembling strategies (Wang et al., 2022a). Illustrative examples of prompt design strategies are provided in Appendix A.1.

This section provides an introduction to the settings of the zero-shot prompt and few-shot prompt, along with a comprehensive description of our prompt definition. A detailed analysis of the GPT-4V’s generated reports under various prompts will be presented in Section 3.2.

**Zero-shot Prompt Scenario** The zero-shot prompt is employed to assess GPT-4V’s capacity to autonomously generate reports without external guidance. To facilitate a comprehensive comparison with the Ground Truth report, we tasked GPT-4V with generating both the expression and findings sections.

**Few-Shot Prompts Scenario** In-context few-shot learning represents a crucial methodology for enhancing the capabilities of large language models (Tsimpoukelli et al., 2021; Wei et al., 2022a; Dai et al., 2022). It enables the model to acquire the necessary output format by providing a set of examples. In contrast to fine-tuning, this method empowers the model to generate desired results without any parameter updating at inference time. We evaluated the in-context few-shot learning capability of GPT-4V using diverse prompt examples. Within the scope of our evaluation, we employ contextual learning to facilitate GPT-4V in generating responses that closely align with the form of ground truth, facilitating meaningful comparisons.

In our investigation of few-shot prompts for the GPT-4V, we conducted experiments with a range of prompt strategies designed for GPT-4V. Specifically, we explored diverse compositions:

- Exclusively using normal examples **(Few-shot normal examples prompt)**;
- Exclusively using abnormal examples **(Few-shot abnormal examples prompt)**;
- Combining one normal and one abnormal example **(Few-shot mixed examples prompt)**;

The details of example reports in prompts are shown in Appendix A.1. Our observations highlighted the substantial impact of prompt type on the model’s output. Depending on the chosen prompt, the model displayed a clear preference either for generating normal reports or abnormal reports. Details will be discussed in section 3.2.

#### 3.1.4 Comparison with Sota

Table 3 presents a comprehensive performance comparison between the GPT-4V model and state-of-the-art methods using the MIMIC-CXR dataset (Johnson et al., 2019). The methods encompasses standard image captioning techniques, including Show-Tell (Vinyals et al., 2015), Att2in (Xu et al., 2015), AdaAtt (Lu et al., 2017), Transformer (Vaswani et al., 2017), and M2Transformer (Cornia et al., 2020). Additionally, the evaluation methods also have medical report generation methods, specifically R2Gen (Chen et al., 2020), R2GenCMN Chen et al. (2021), MSAT (Wang et al., 2022b), and METransformer (Wang et al., 2023a). To provide fair comparisons, we employ the exact same prompting structure **(Few-shot mixed examples prompt)** to help GPT-4V generate the medical report.

**Table 1:** Comparison MIMIC-CXR datasets.

| Dataset | Methods | BLEU-1 | BLEU-2 | BLEU-3 | BLEU-4 | ROUGE | METEOR | CIDEr |
| --- | --- | --- | --- | --- | --- | --- | --- | --- |
| MIMIC-CXR | Show-Tell (Vinyals et al., 2015) | 0.308 | 0.190 | 0.125 | 0.088 | 0.256 | 0.122 | 0.096 |
|  | Att2in (Xu et al., 2015) | 0.314 | 0.198 | 0.133 | 0.095 | 0.264 | 0.122 | 0.106 |
|  | AdaAtt (Lu et al., 2017) | 0.314 | 0.198 | 0.132 | 0.094 | 0.267 | 0.128 | 0.131 |
|  | Transformer (Vaswani et al., 2017) | 0.316 | 0.199 | 0.140 | 0.092 | 0.267 | 0.129 | 0.134 |
|  | M2Transformer (Cornia et al., 2020) | 0.332 | 0.210 | 0.142 | 0.101 | 0.264 | 0.134 | 0.142 |
|  | R2Gen (Chen et al., 2020) | 0.353 | 0.218 | 0.145 | 0.103 | 0.277 | 0.142 | - |
|  | R2GenCMN (Chen et al., 2021) | 0.353 | 0.218 | 0.148 | 0.106 | 0.278 | 0.142 | - |
|  | PPKED (Liu et al., 2021) | 0.360 | 0.224 | 0.149 | 0.106 | 0.284 | 0.149 | 0.237 |
|  | GSK (Yang et al., 2021b) | 0.363 | 0.228 | 0.156 | 0.115 | 0.284 | - | 0.203 |
|  | MSAT (Wang et al., 2022b) | 0.373 | 0.235 | 0.162 | 0.120 | 0.282 | 0.143 | 0.299 |
|  | METransformer (Wang et al., 2023a) | 0.386 | 0.250 | 0.169 | 0.124 | 0.291 | 0.152 | 0.362 |
|  | GPT-4V (OpenAI, 2023) | 0.338 | 0.190 | 0.109 | 0.061 | 0.240 | 0.125 | 0.033 |

**Table 2:** Visual Question Answering Method Comparison.

| Dataset | Reference Methods | Fusion Method | Close-end |
| --- | --- | --- | --- |
| VQA-RAD | StAn (He et al., 2020) | SAN | 57.2 |
|  | BiAn (He et al., 2020) | BAN | 67.9 |
|  | MAML (Finn et al., 2017) | SAN | 69.7 |
|  |  | BAN | 72.4 |
|  | MEVF (Nguyen et al., 2019) | SAN | 74.1 |
|  |  | BAN | 75.1 |
|  | MMQ (Do et al., 2021) | SAN | 75.7 |
|  |  | BAN | 75.8 |
|  | PubMedCLIP (Eslami et al., 2021) | - | 80 |
|  | MMBERT (Khare et al., 2021) | - | 77.9 |
|  | Q2ATransformer (Liu et al., 2023) | - | 81.2 |
|  | GPT-4V (OpenAI, 2023) | - | 61.40 |

**Table 3:**
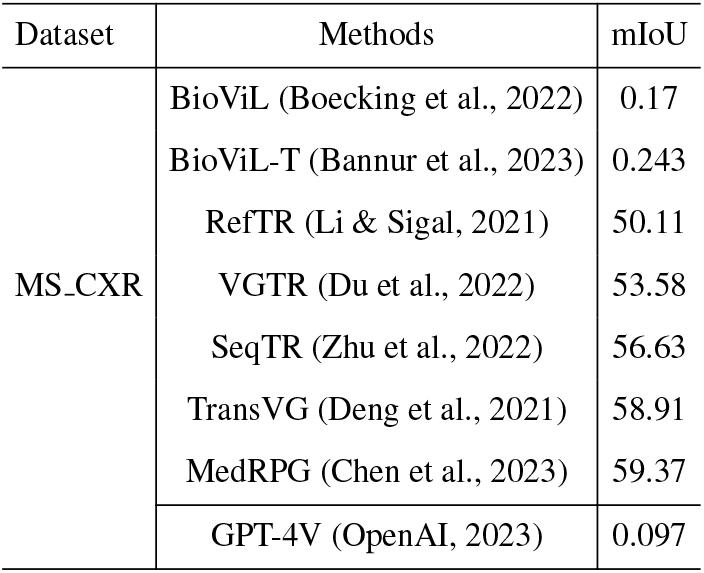
Comparison MS-CXR datasets.

| Dataset | Methods | mIoU |
| --- | --- | --- |
| MS-CXR | BioViL (Boecking et al., 2022) | 0.17 |
|  | BioViL-T (Bannur et al., 2023) | 0.243 |
|  | RefTR (Li & Sigal, 2021) | 50.11 |
|  | VGTR (Du et al., 2022) | 53.58 |
|  | SeqTR (Zhu et al., 2022) | 56.63 |
|  | TransVG (Deng et al., 2021) | 58.91 |
|  | MedRPG (Chen et al., 2023) | 59.37 |
|  | GPT-4V (OpenAI, 2023) | 0.097 |

From Table 3, it’s clear that medical report generation models such as METransformer, MSAT, and R2Gen showcase top-tier performance. Nevertheless, GPT-4V’s capability to generate medical reports is impressive, even if it’s a general-purpose model. Leveraging the advantages of an extensive pre-training dataset, it excels in several metrics, including BLEU, ROUGE, and METEOR. However, when compared to models specifically trained on MIMIC-CXR, it exhibits a gap, particularly evident in the CIDEr metric. This discrepancy arises because the CIDEr metric assigns varying score weights to words based on their occurrence frequencies, potentially limiting GPT-4V’s performance in generating certain MIMIC-CXR-specific words, consequently yielding relatively lower scores.

Furthermore, our testing has revealed that GPT-4V possesses the capacity to generate information that is absent in the ground truth but is visually evident in the image. This phenomenon contributes to GPT-4V’s relatively lower performance on metrics such as BLEU, which primarily assesses word match rates. One example is shown in Figure 1.

**Figure 1:**
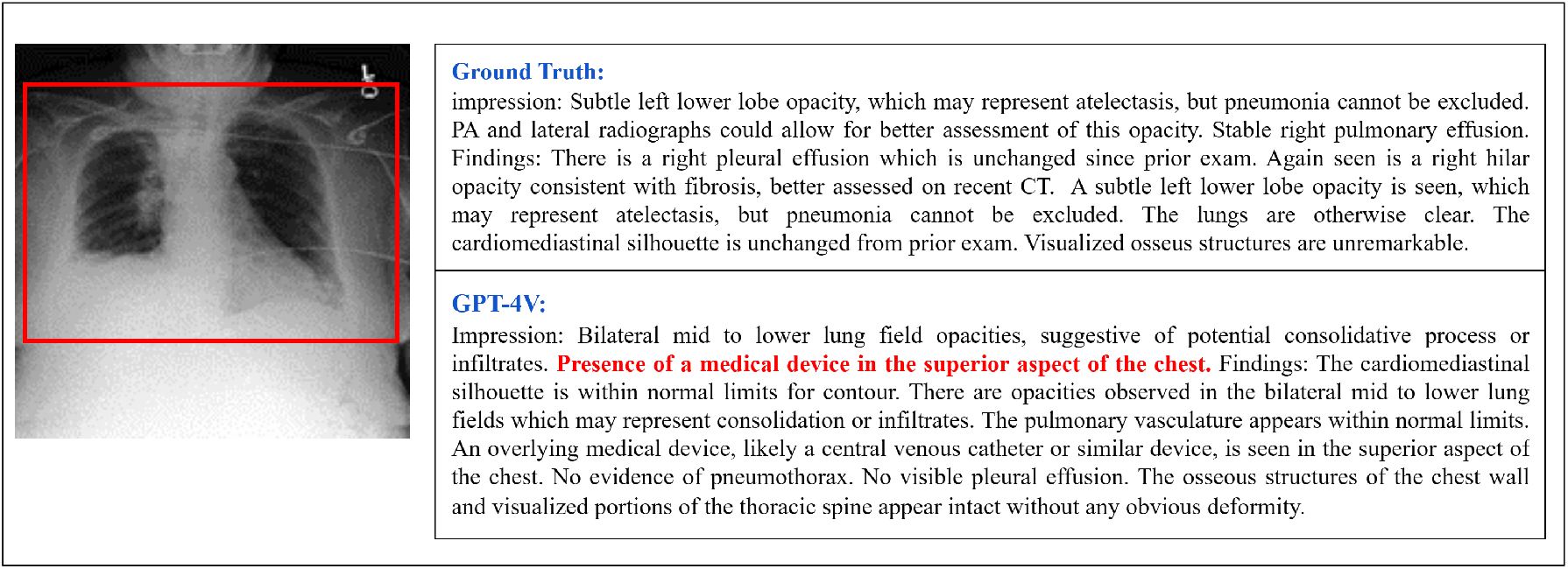
One case with few-shot mixed examples prompt. The ground truth does not reference a medical device; however, one is visibly present in the image and indicated by a red box. GPT-4V demonstrates the ability to recognize this medical device.

### 3.2 Case Study

#### 3.2.1 Zero-shot Behavior

In the zero-shot scenario, through a series of tests on multiple chest X-ray images, we observed that GPT-4V consistently generates reports with a focus on various anatomical organs. This phenomenon is illustrated in Figure 11. Notably, GPT-4V tends to follow a specific order, including the information of lung, cardiomediastinal silhouette, bones, diaphragm, and soft tissues, for the majority of the generated reports.

While the format of the generated reports may vary from MIMIC-CXR, the content within these reports does convey both normal and abnormal aspects of the radiographic images. Figure 2 shows a selection of examples. The observations reveal that GPT-4V can describe the normal aspects in the images. Furthermore, as demonstrated in Example 3, GPT-4V exhibits the capacity to recognize abnormalities, *’suggestive of a possible infectious or inflammatory process’*. These instances collectively underscore that, even in the context of Zero-shot prompts, GPT-4V may not replicate the exact report format found in MIMIC-CXR, yet it demonstrates a noteworthy ability to generate relevant reports and identify anomalies.

**Figure 2:**
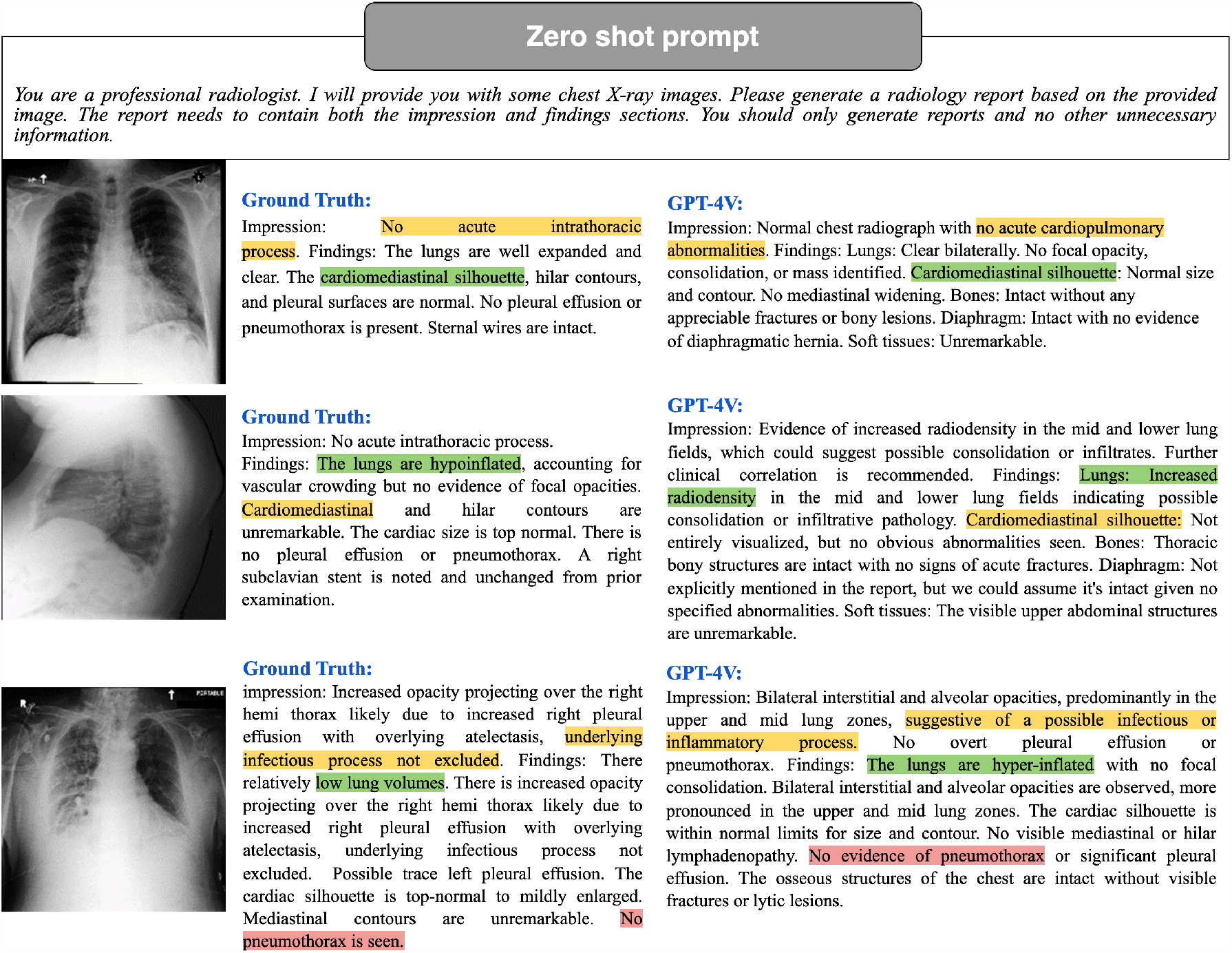
Zero-shot prompt example. GPT-4V can generate radiology reports without example reports and can convey both normal and abonrmal aspects. For better illustration, the key medical information in the reports is highlighted using different colors.

#### 3.2.2 Few-shot Behavior

In this prompt scenario, we explored 3 kinds of prompt settings:

- **Few-shot normal examples prompt**
- **Few-shot abnormal examples prompt**
- **Few-shot mixed examples prompt**

In this section, we present a comprehensive analysis of reports generated by GPT-4V under three distinct few-shot prompts. We observe that different prompts significantly influence the generated reports. Specifically, Figure 3 illustrates the response to a normal chest X-ray image, where we employ three distinct prompt methodologies to guide GPT-4V in generating corresponding reports. Interestingly, the reports generated from the normal examples prompt and mixed examples prompt both describe the image as normal. In contrast, the report originating from the abnormal examples prompt highlights anomalies. This indicates that GPT-4V’s inclination to generate normal or abnormal reports varies based on the provided example reports.

**Figure 3:**
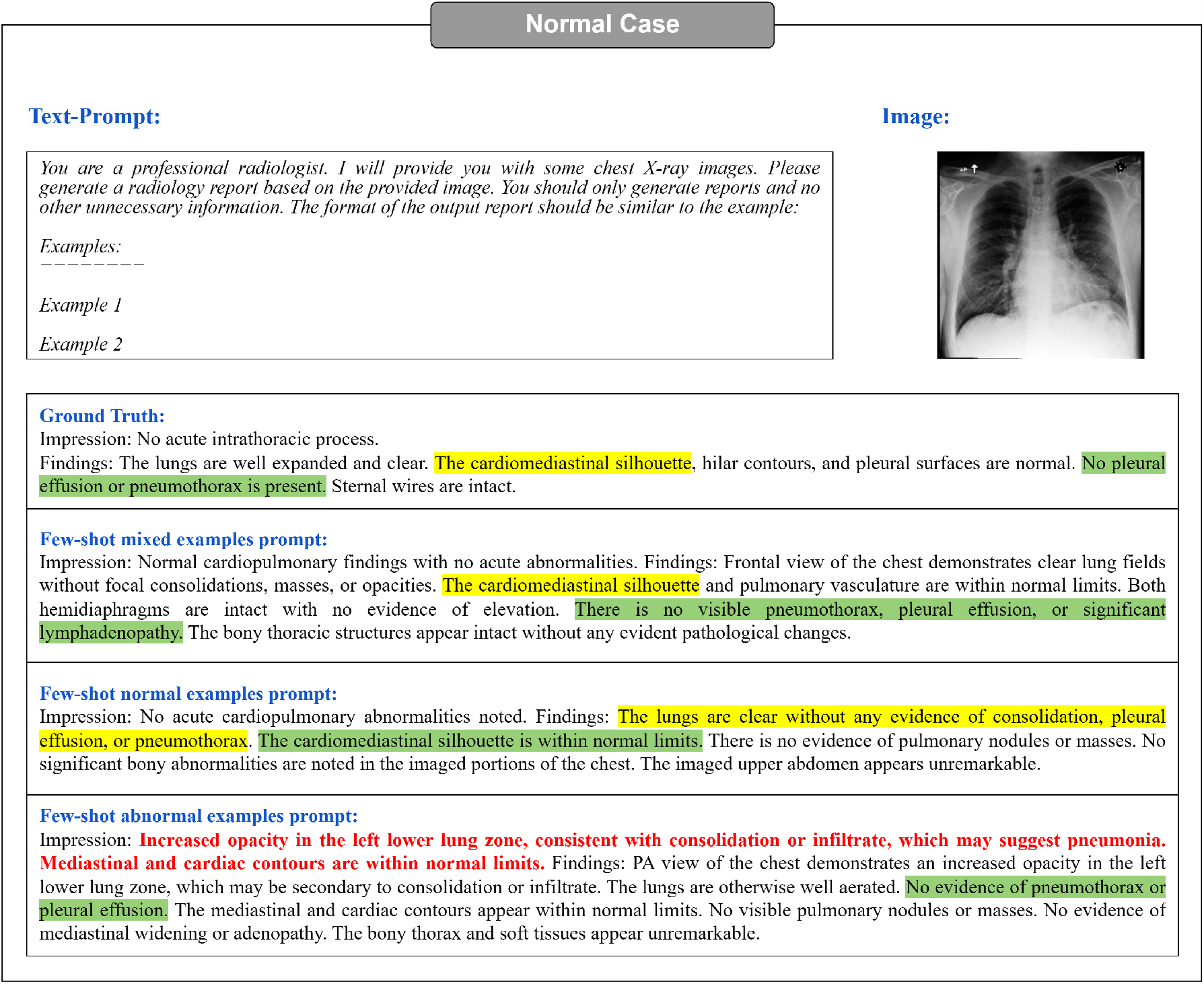
Few-shot normal case (The key medical information in the reports is highlighted using different colors). GPT-4V is more likely to generate abnormal reports when the prompt includes two abnormal examples. The words in red correspond to descriptions of abnormal conditions.

The analysis of reports generated for an abnormal chest X-ray image can be found in the appendix A.2 with a more detailed explanation. However, it’s worth noting here that our subsequent tests have shown that the mixed examples prompt (illustrated in figure 3, 14) has a significant influence on GPT-4V’s capacity to accurately determine the normalcy or abnormality of an image. Due to this observed consistency and reliability, we opted for the mixed examples prompt when testing the entire MIMIC-CXR test set and in the computation of related evaluation metrics.

For these examples, we can know summarize the impact of different prompts on the generated reports as follows:

**Normal Examples Prompt** The generated report focuses on the normal aspects of the image, seemingly overlooking or not emphasizing the abnormalities present. This could be attributed to the inherent bias introduced by the normal examples in the prompt, steering the GPT-4V’s focus towards more routine or standard interpretations.

**Abnormal Examples Prompt** As expected, the report provides a clear and distinct description of the abnormalities evident in the X-ray. However, for normal chest X-ray radiographs, the GPT-4V may also exhibit a heightened probability of generating certain erroneous indications of abnormality.

**Mixed Examples Prompt** The mixed examples prompt leads the GPT-4V to accurately describe the abnormal and normal conditions of the image. This suggests a balanced effect, where the GPT-4V doesn’t get overly biased by either the normal or abnormal examples but leverages both to arrive at an accurate interpretation.

From this in-depth examination, it becomes evident that the choice of prompt plays a pivotal role in guiding GPT-4V’s performance, especially when anomalies are present in medical images. The mixed examples prompt, in particular, shows promise in achieving a balanced and accurate report, making it a potential choice for diverse medical scenarios.

#### 3.2.2 Prompt Augmentation for Output view information

Additionally, our investigations revealed that augmenting the information content within a given prompt enables GPT-4V to produce more pertinent information in its generated reports. As an illustrative example, we incorporated instances with view information for chest X-ray images within both the few-shot mixed examples prompt and the few-shot abnormal Examples prompt. Conversely, view information was omitted in the few-shot normal examples prompt. This deliberate contrast in prompt content demonstrated that prompts containing view information effectively instructed GPT-4V to incorporate considerations of image viewpoint into the report generation process.

More specifically, we supplemented the few-shot mixed examples prompt and the few-shots abnormal examples prompt with the following view information:

- Frontal and lateral views of the chest;
- PA and lateral views of the chest provided.

As illustrated in Figure 4,15 the inclusion of view information prompts GPT-4V to incorporate corresponding viewpoint details into the generated report. For instance, it generates phrases like *’PA view of the chest provided’* and *’Frontal view of the chest demonstrates*…*’*. However, it is essential to acknowledge that while enhancing the prompt with view information empowers GPT-4V to produce reports enriched with these details, there are instances where GPT-4V inaccurately identifies viewpoint information. The incorrect case is shown in Appendix A.3.

**Figure 4:**
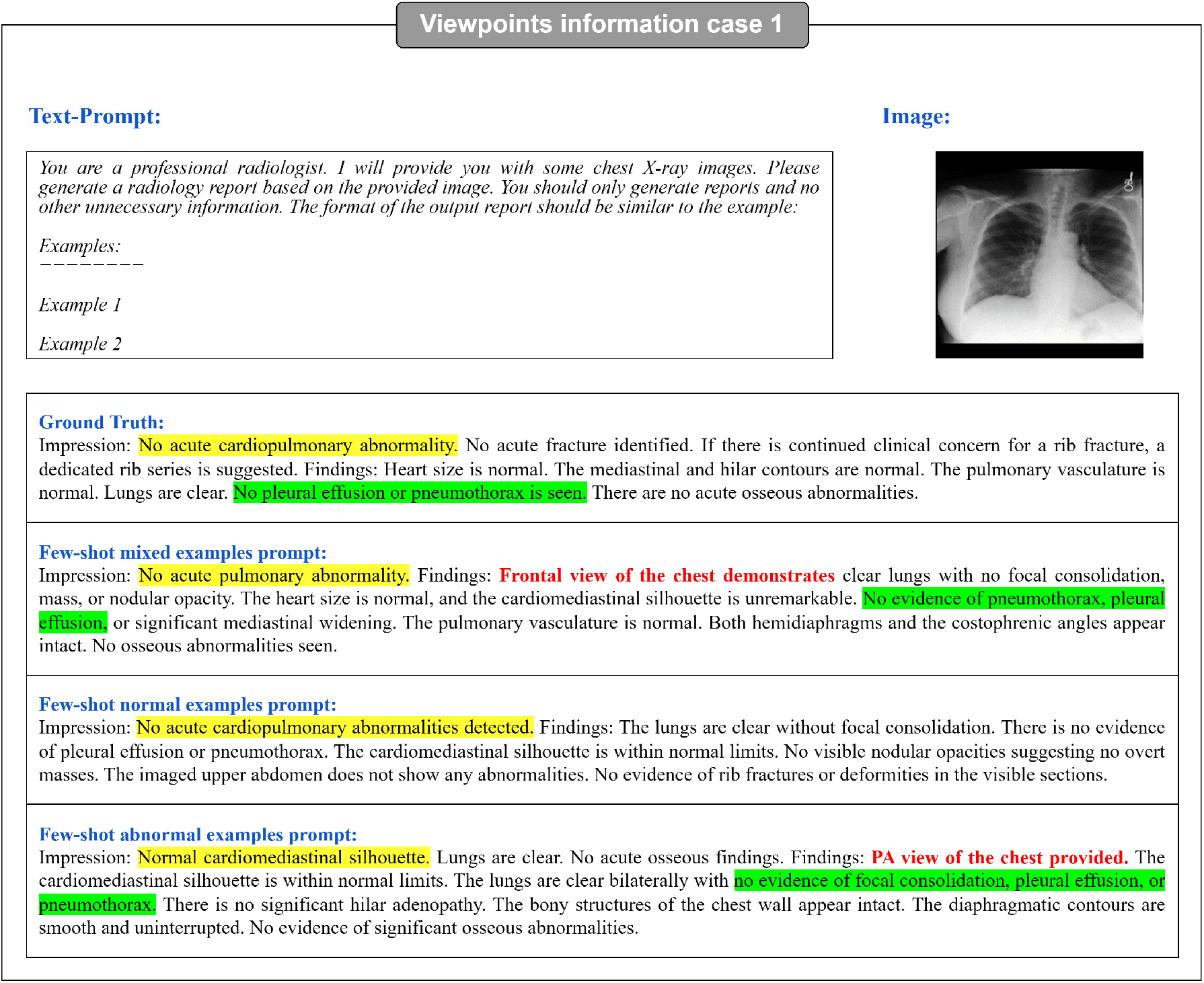
Viewpoint information Case 1 (The key medical information in the reports is highlighted using different colors). The inclusion of view information in the prompt results in a higher probability of GPT-4V generating view information, indicated in red text in the figure. Notably, GPT-4V does not generate view information when the prompt lacks such information, as seen in the normal examples prompt (in Figure 13).

This phenomenon can be attributed to two primary factors: firstly, potential constraints in GPT-4V’s inherent recognition capabilities, and secondly, the potential inadequacy of prompt design in fully activating GPT-4V’s ability to discern viewpoint information.

It is imperative to emphasize that, even with the incorporation of view information, the core content within the generated reports exhibits a high degree of consistency (crucial medical information in the reports is distinguished using diverse colours in Figure 4). This observation leads to a significant conclusion: the inclusion of supplementary information within the prompt broadens the spectrum of content integrated into the generated report, all while preserving GPT-4V’s capability to fulfill common tasks.

These examples vividly illustrate the critical role of prompt design within the domain of in-context few-shot learning. In contrast to the fine-tuning approach, few-shot learning empowers GPT-4V to gain essential knowledge from the prompt and subsequently apply this knowledge in generative tasks. Consequently, the meticulous design of a logical and effective prompt emerges as a pivotal factor when leveraging GPT-4V for medical report generation tasks. This aspect of prompt design deserves future studies.

### 3.3 Discussion

Our extensive evaluation and case study of GPT-4V’s capabilities in Radiology Report Generation reveal its potential as well as its current limitations. By employing various prompts, GPT-4V demonstrates the capacity to generate descriptive reports for chest X-ray images, covering both normal and abnormal aspects. Remarkably, the design of the prompt significantly influences GPT-4V’s performance; prompts with more information lead to greater attention to the image and the generation of more detailed descriptions.

It is essential to highlight that GPT-4V was not trained specifically on MIMIC-CXR, which impacts its capacity to generate specific rare words, leading to relatively lower scores on commonly used evaluation metrics. Nevertheless, GPT-4V demonstrates the ability to generate content related to images that is not explicitly mentioned in the Ground Truth but is visually apparent. As a result, further research aimed at improving GPT-4V’s report accuracy remains a valuable pursuit.

## 4 Medical Visual Question Answering

Visual Question Answering (VQA) has become a much critical research area. The goal of VQA systems is to enable computers to understand natural language questions and provide accurate answers on images. In the following, we will explore the medical image VQA performance of GPT-4V on the VQA-RAD dataset and compare it with the current SOTA method.

### 4.1 Evaluation

In order to assess GPT-4V’s effectiveness on the Medical VQA dataset, we embarked on a comprehensive series of experiments. Utilizing the GPT-4V model, we applied it to generate predicted answers based on the input medical image and the question related to this image. Then, proceeded to calculate the accuracy of the results. Subsequently, we conducted a comparative analysis with the current state-of-the-art (SOTA) methods. Herein, we present our principal observations and conclusions.

#### 4.1.1 Dataset: VQA-RAD

VQA-RAD (Lau et al., 2018) is one of the most widely utilized radiology datasets. It comprises 315 images along with 3515 question-answer pairs, ensuring that each image corresponds to at least one question-answer pair. The questions encompass 11 distinct categories, including “anomalies,” “properties,” “color,” “number,” “morphology,” “organ type,” “other,” and “section.” A noteworthy 58% of these questions are designed as closed-ended queries, while the remainder take the form of open-ended inquiries. These images predominantly feature the head, chest, and abdomen regions of the human body. It is essential to manually partition the dataset into training and test sets for accurate evaluation.

#### 4.1.2 Overview of Prompt Methods

GPT-4V not only possesses powerful natural language processing capabilities, but also incorporates advanced computer vision techniques, which makes it excel in handling fusion tasks of images and text. It is trained to understand questions and extract information from images to generate accurate answers. However, the performance of GPT also depends on how the design of the prompt.

To ensure that GPT-4V accurately grasps the answer style of the VQA-RAD dataset, we provided seven examples to guide the model in generating responses consistent with the dataset’s format. Without these examples, GPT-4V tends to produce more unconstrained answer text, complicating the task of comparing predicted answers with the ground truth.

We Designed the prompt by following the template in Figure 5:

**Figure 5:**
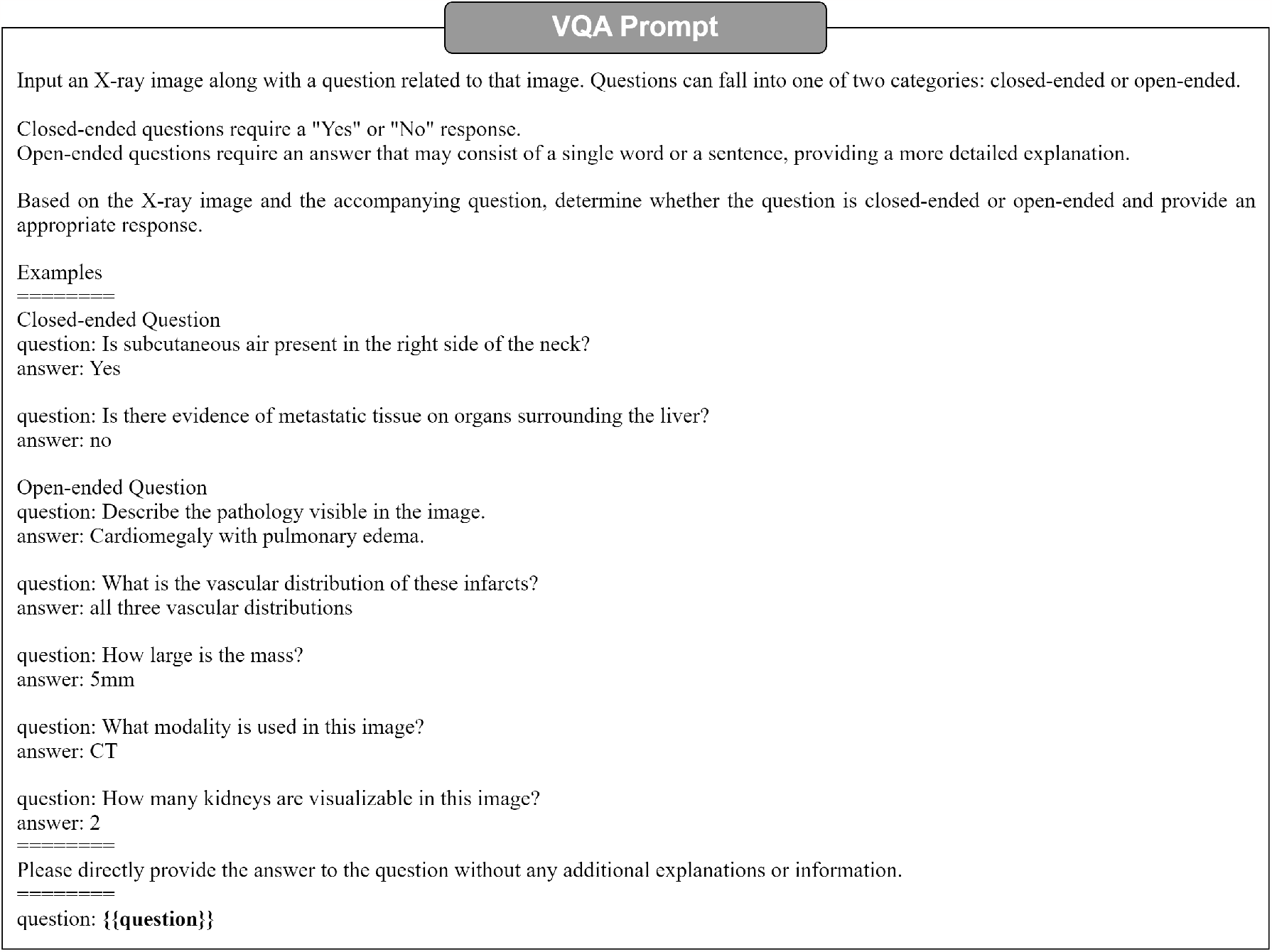
VQA Prompt Method. Elements in double braces are replaced with specific questions

#### 4.1.3 Comparison with SOTA

Upon scrutinizing the results of GPT-4V on the VQA-RAD dataset’s test set, it is calculated that the accuracy for closed-end questions is 61.4%, the result shows in table2, which is significantly lower than other published results. In terms of open-end questions, the calculated BLEU score is 0.1155, which also does not reach a high standard. The majority of currently available research primarily employs classification based model to tackle Visual Question Answering (VQA) problems. This approach results in a lack of evaluations using the BLEU score, making it challenging to draw comparisons between different methods. However, upon analyzing the cases provided by GPT-4V, it is postulated that the low BLEU score may be attributed to the excessive flexibility of GPT-4V, resulting in substantial deviations from the correct answers. This might be due to some clear limitations of BLEU itself. BLEU lacks semantic understanding, as it mainly relies on the literal matching of n-grams and does not deeply understand context and semantics. It is insensitive to synonyms and diverse ways of expression. Even if two sentences mean the same thing, if they use different words or ways of expression, the BLEU score might end up being quite low. In simpler terms, BLEU struggles to recognize when different words mean the same thing, and this can lead to unfairly low scores even when the answers are correct. We hope that in the future, more advanced methods capable of deeply understanding the semantics of text will be developed, providing more accurate and reliable assessments.

### 4.2 Case Study

We present few case study of VQA in Figure 6 7 From the case study, we can tell that the GPT-4V showed some limitations in the Medical VQA domain. It showed strong ability in determining whether a question was close-end or open-end, and was almost always able to make a correct judgment. However, in answering some open-end questions, it did not make full use of the image information, relying instead on the medical terms mentioned in the question itself, and failing to make effective reference to the information in the medical images. For example, in the last case, the GPT-4V only expanded on the nouns that appeared in the question without taking the medical images into account, resulting in an incorrect answer. There were also some instances of incorrect responses to close-end questions. These questions did not perform as well as expected, and further improvements and optimizations are needed to improve performance in Medical VQA tasks.

**Figure 6:**
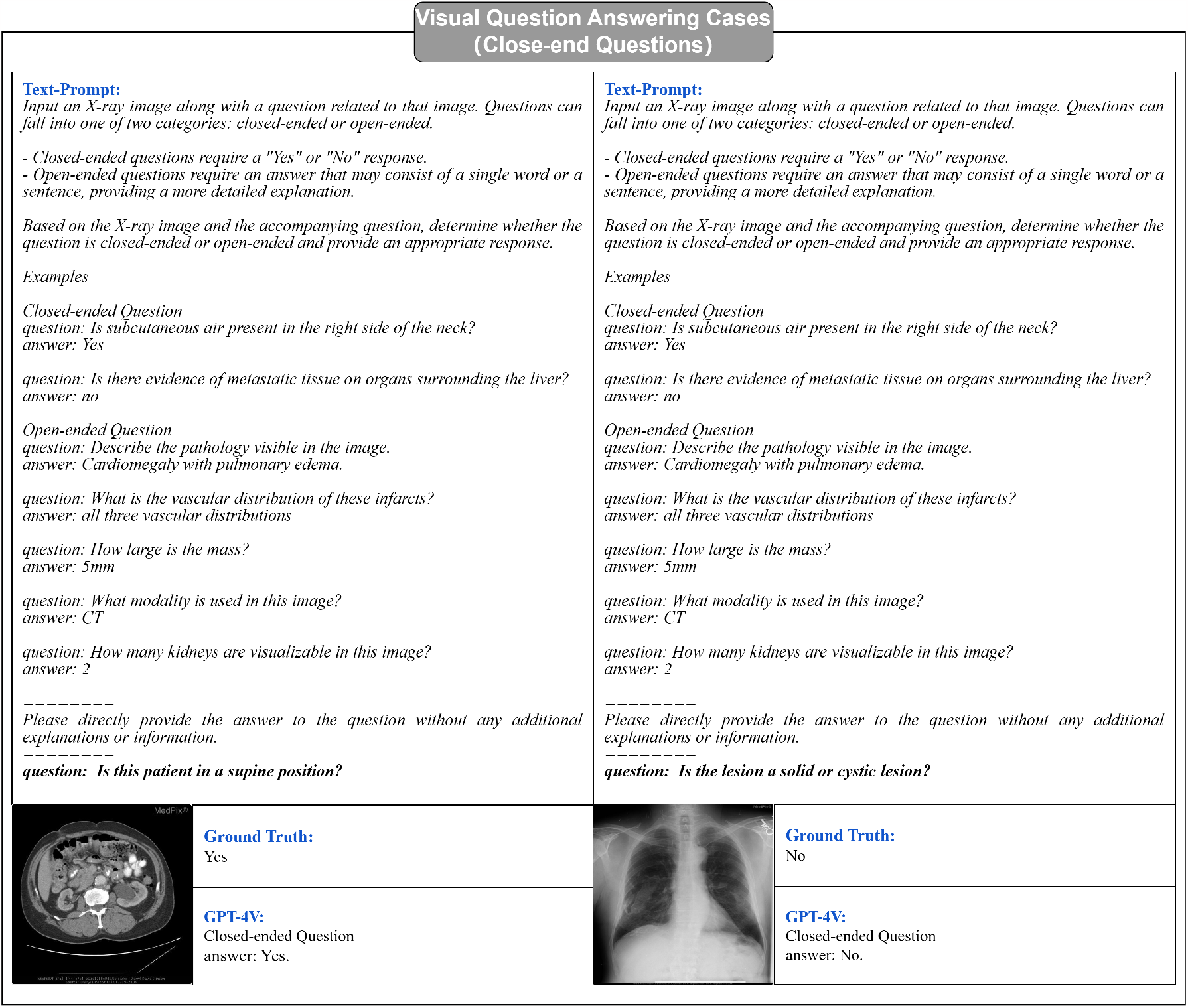
VQA Prompt examples.By given few-shot prompts, GPT-4V can generate answers for the given image and question pairs, the result for the close-end question is better than open-end questions

**Figure 7:**
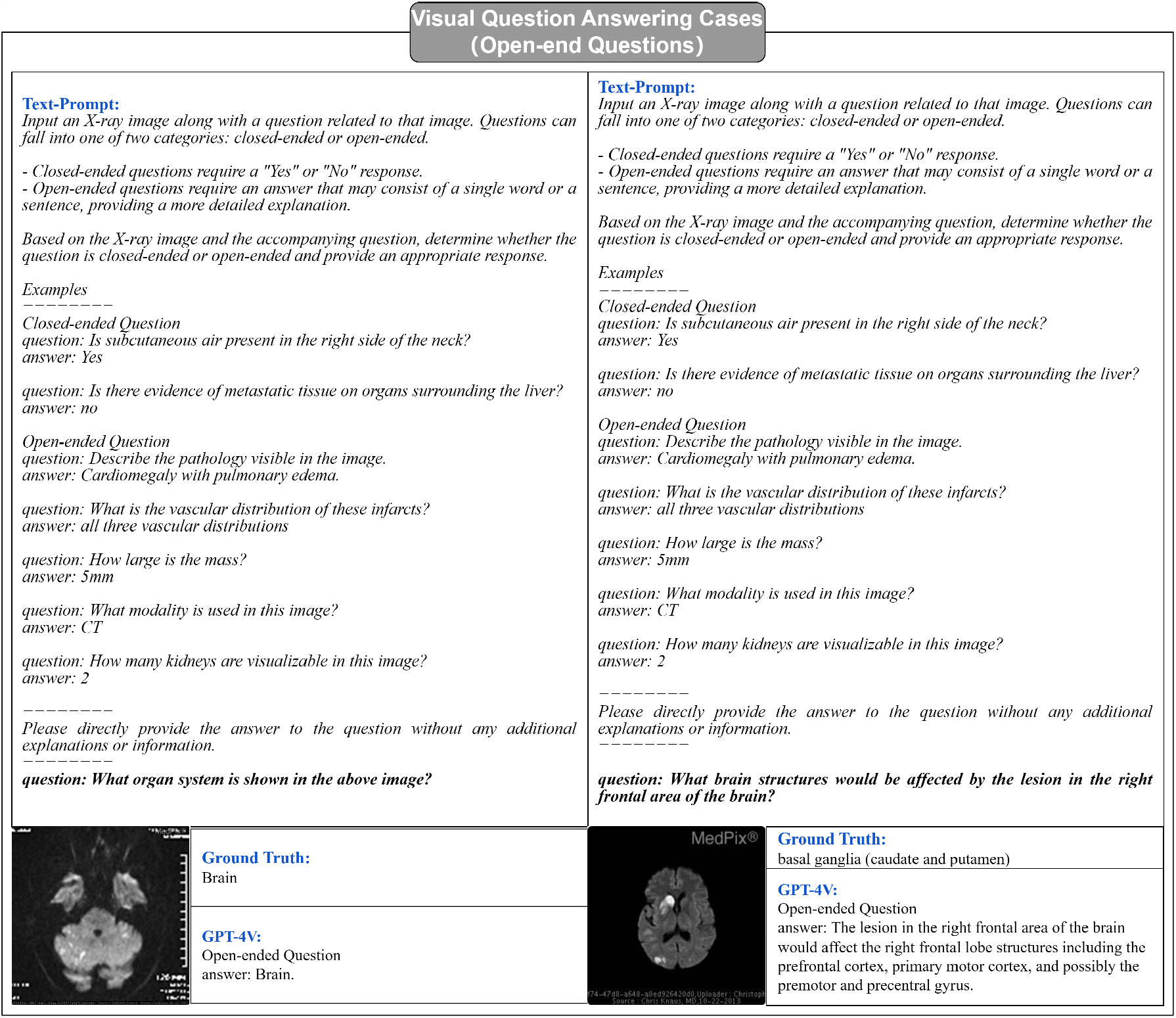
VQA Prompt examples. With the assistance of a few-shot prompts, GPT-4V has the capability to generate responses for open-ended questions, though there is room for refinement to enhance its performance.

### 4.3 Discussion

Our extensive evaluation and in-depth case studies of GPT-4V’s performance on the VQA-RAD dataset have highlighted its potential capabilities as well as the areas that necessitate substantial improvement within the Medical Visual Question Answering (VQA) field.

While GPT-4V demonstrates proficiency in distinguishing between closed-end and open-end questions, its accuracy rate of 61.4% for closed-end questions and low BLEU score of 0.1155 for open-end questions signify a performance level that is considerably below the published benchmarks in this domain. This discrepancy underscores the need for more refined and optimized models that can more accurately interpret and respond to medical imagery. The capability to accurately identify whether a question is open-ended or closed-ended demonstrates GPT’s substantial reasoning skills. However, its occasional low accuracy could be attributed to an insufficient amount of training data. Acquiring general Visual Question Answering (VQA) data is relatively easier compared to procuring medical VQA data. This discrepancy is due to the labor-intensive and expensive nature of labeling medical data. Consequently, as the volume of training data in the medical domain increases, we can anticipate an enhancement in the performance of VQA applications.

Furthermore, the limitations of the BLEU score as an evaluation metric, particularly its lack of semantic understanding and sensitivity to diverse expressions and synonyms, have been highlighted. This brings to light the urgent need for the development of more advanced and semantically aware evaluation methods to provide accurate and reliable assessments of model performance in this field.

## 5 Medical Visual Grounding

Visual Grounding is one of the important tasks in the field of computer vision, aimed at enabling computers to understand natural language descriptions and associate them with specific regions in an image. This technique has great potential in areas such as medical image analysis. In this paper, we presented the performance of GPT-4V on MS-CXR dataset for visual grounding applications and compare it with current SOTA methods.

### 5.1 Evaluation

#### 5.1.1 Dataset: MS-CXR

The MS-CXR (Boecking et al., 2022) dataset is a valuable resource for biomedical vision-language processing, featuring 1162 image-sentence pairs with bounding boxes and corresponding phrases. It was meticulously annotated by board-certified radiologists, covering eight cardiopulmonary radiological findings, each having an approximately equal number of pairs. This dataset offers both reviewed and edited bounding boxes/phrases and manually created bounding box labels from scratch. What sets MS-CXR apart is its focus on complex semantic modeling and real-world language understanding, challenging models with joint image-text reasoning and tasks like parsing domain-specific location references, complex negations, and variations in reporting style. It serves as a benchmark for phrase grounding and has been instrumental in demonstrating the effectiveness of principled textual semantic modeling for enhancing self-supervised vision-language processing.

#### 5.1.2 Overview of Prompt Methods

We’ve looked at many different ways to give instructions to GPT, and we’ve found a specific type that helps it understand better and makes it easier to create bounding boxes. We chose this prompt after carefully checking which one work best. We Designed the prompt by following the template in Figure 8:

**Figure 8:**
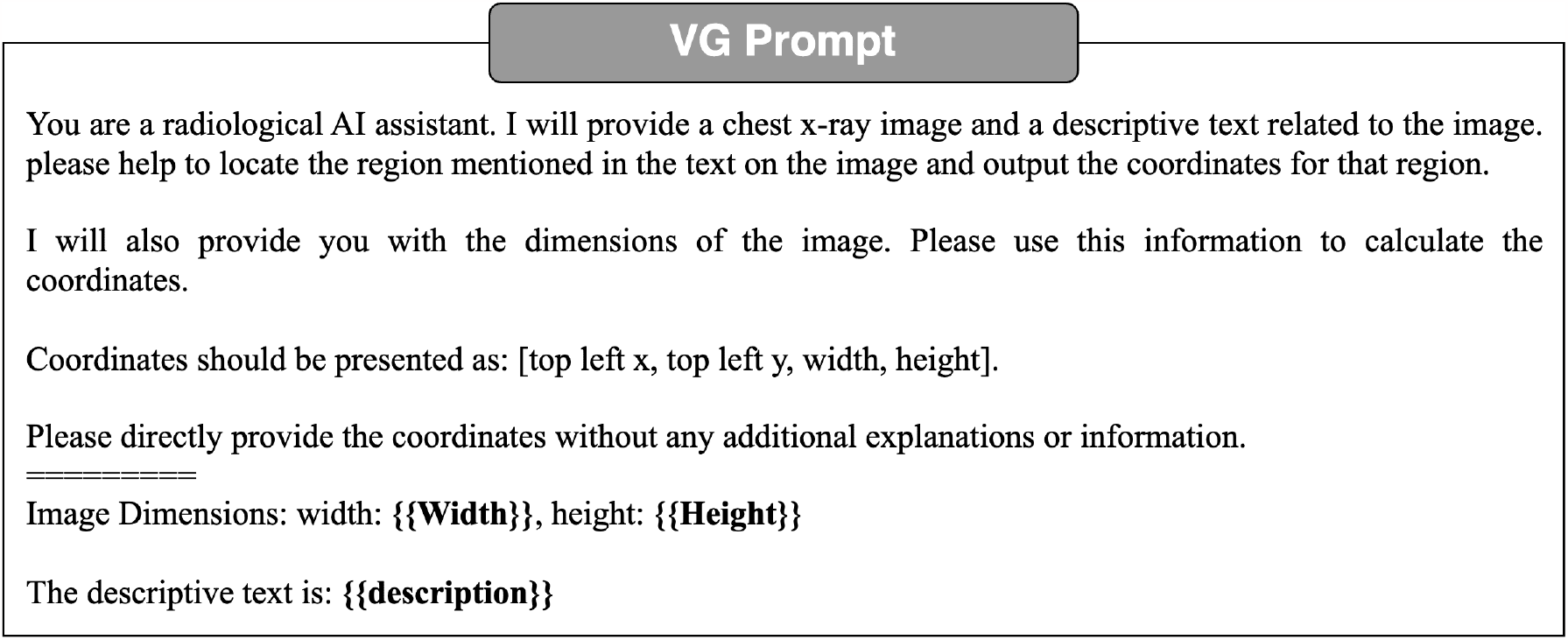
VG Prompt Method. Elements in double braces are replaced with specific image width, height and description text related to image

**Figure 9:**
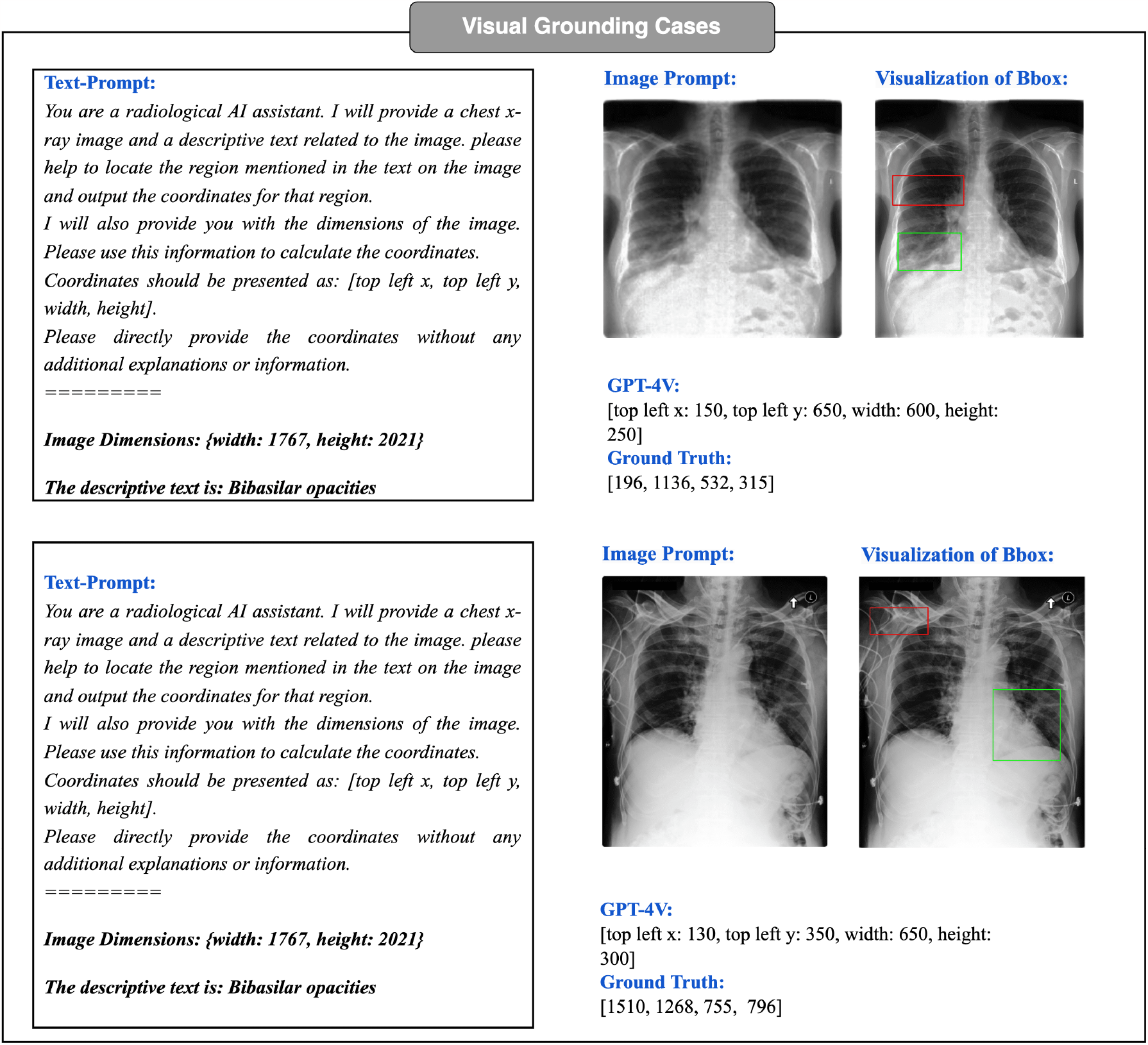
Visual Grounding Prompt examples. The bounding boxes in red color are predicted box by GPT-4V, and the green bounding boxes are ground truth boxes. GPT-4 is capable of generating and estimating the bounding box coordinates for the reference text within an image. However, the results show that the GPT-4V can not understand medical image properly.

**Figure 10:**
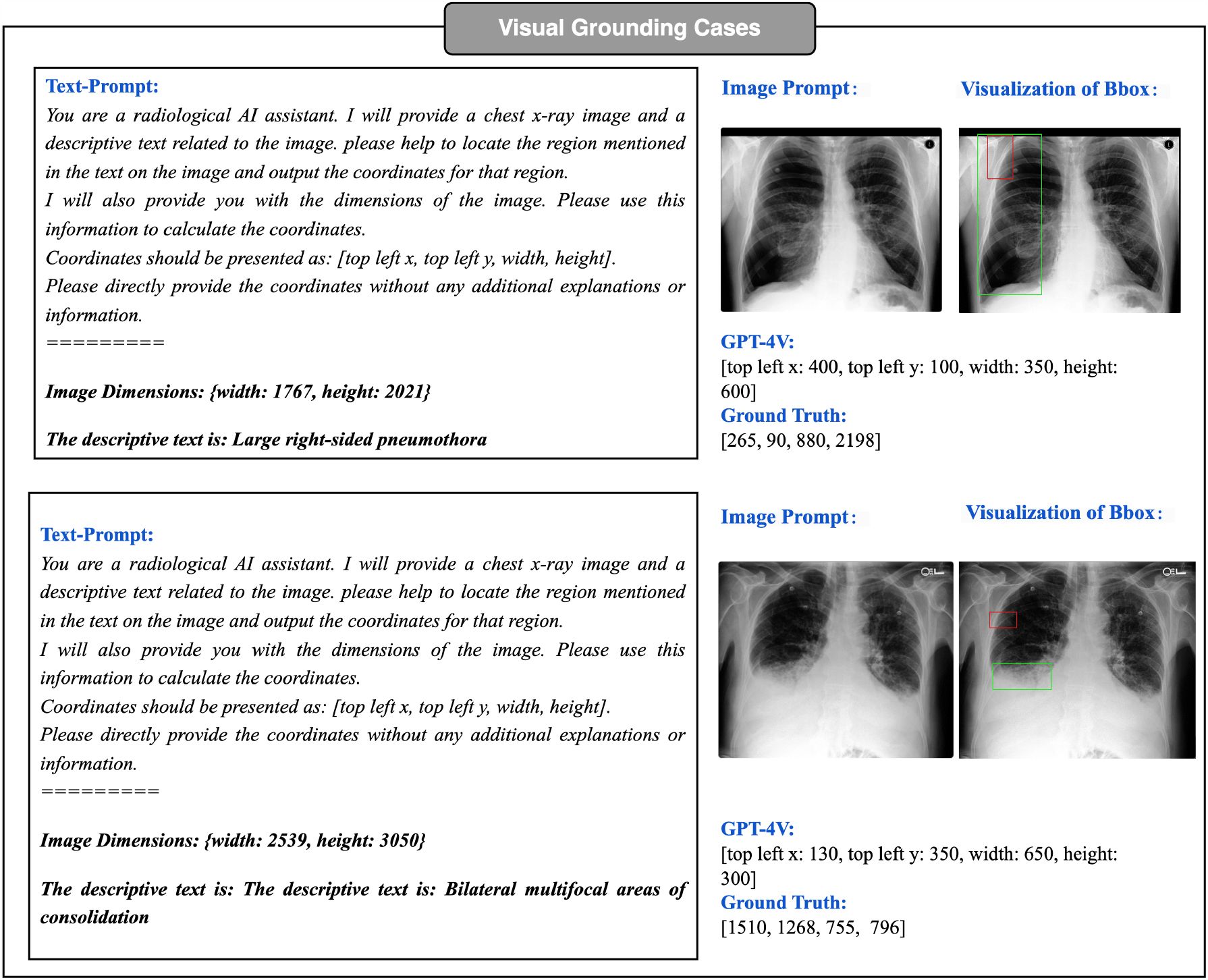
There are some Visual Grounding Prompt examples. The bounding boxes in red color are predicted box by GPT-4V, and the green bounding boxes are ground truth boxes.

#### 5.1.3 Comparison with SOTA

In order to compare with current existing models, we use mean Intersection-Over-Union(mIoU) as our evaluation metrics. Upon conducting an evaluation of GPT-4V’s performance on the MS-CXR dataset, the calculated mean Intersection over Union (mIoU) was found to be 0.0974. This result is markedly lower than all published benchmarks. Empirical evidence demonstrates that while GPT-4V possesses the capability to comprehend applications within Visual Grounding, it exhibits a deficiency in accurately identifying medical organs and pathological signs. Consequently, this results in imprecise bounding box predictions. Recently, SoM (Yang et al., 2023a) addressed this issue and made significant improvements. The approach in the paper involved first segmenting and labeling the image, and then proceeding with grounding, which led to substantial enhancements in performance. However, this method was applied to general images, and it’s not certain that it would yield equally impressive results for medical images, which require much finer-grained features. Further experiments will be necessary to validate its effectiveness in such contexts.

### 5.2 Case study

From the case study, it appears that the GPT-4V has the potential to generate bounding boxes, but notably, its accuracy performs rather poorly. Although it was able to attempt to calibrate the position of the object, there were serious errors and uncertainties in this task. This may be due to the fact that GPT-4V’s model has some limitations in processing the image information and is unable to fully understand and interpret the exact position of the object in the image. Especially for the medical image, which need more focus on fine grain feature. Another possible reason for GPT-4V’s poor performance could be that it was mainly trained using common, everyday images, and it didn’t have a lot of varied images to learn from. The GPT model needs a lot more data to work well and become reliable. So, because it didn’t have enough diverse data to learn from, it doesn’t perform very well.

### 5.3 Discussion

Our comprehensive evaluation and case study of GPT-4V’s capabilities in Visual Grounding highlight both its potential and its current limitations. While the model shows promise in recognizing bounding boxes, it falls significantly short in terms of accuracy, as evidenced by its low mIoU score when compared to existing benchmarks and its performance on the MS-CXR dataset. Its inability to precisely identify medical organs and pathological signs leading to imprecise bounding box predictions, and this may caused by lack of training data. It is very hard to get enough labeled data for GPT to train.

In light of these findings, it is evident that GPT-4V requires further refinement and training to overcome its current limitations and to enhance its bounding box localization accuracy. In order to achieve better results in this area, further model improvement and more data is needed to increase the accuracy of its bounding box localization, thus making it more useful and reliable in various applications.

Doing so will undoubtedly make GPT-4V a more reliable and valuable tool in various applications, fostering its integration and utility in practical, real-world scenarios, especially within the medical field. This journey towards improvement is not only necessary but also a crucial step in advancing the field of Visual Grounding and in unlocking the full potential of models like GPT-4V.

## 6 Conclusion

The comprehensive assessment of GPT-4V’s capabilities in Radiology Report Generation, Medical Visual Question Answering (VQA), and Visual Grounding offers a perspective on the model’s potential and areas for improvement within the medical domain. GPT-4V’s ability to generate radiology reports based on chest X-ray images is commendable, particularly when furnished with detailed prompts. This underscores the capacity of language models to aid in radiology diagnosis. Nevertheless, its challenges in recognizing uncommon terms and subtle differences specific to the MIMIC-CXR dataset underscore the necessity for domain-specific training and fine-tuning to elevate its proficiency in medical reporting.

Furthermore, although GPT-4V displays proficiency in distinguishing among various question types within the VQA-RAD dataset, its performance metrics, especially for open-ended questions, fall short of public benchmarks. This sub-optimal performance reveals a gap in its comprehension and response capabilities related to medical imaging. Moreover, the limitations of current evaluation metrics like the BLEU score underscore the significance of constructing semantically-aware evaluation methodologies to gain a holistic comprehension of the model’s aptitude.

The Visual Grounding evaluation further explored the difficulties GPT-4V encounters in achieving high precision in bounding box localization within medical images. These limitations, particularly its struggles in identifying medical organs and pathological indicators, underscore the urgent requirement for specialized training and model improvements to enhance its grounding capabilities.

In summary, GPT-4V demonstrates remarkable potential across various medical image analysis domains. Nonetheless, its current limitations underscore the necessity for domain-specific enhancements. Exploring dedicated training on medical datasets, designing comprehensive prompt methodologies, and advancing evaluation techniques still need further research.

## Data Availability

All data produced are available online at https://physionet.org/content/mimic-cxr/2.0.0/.

## A Appendix

### A.1 Details of Prompt Settings

In all prompts, we prompt GPT-4V to assume the role of a professional radiologist. Additionally, we explicitly instruct it to generate both the impression and findings sections.

#### A.1.1 Zero-shot prompt

Figure 11 showcases a zero-shot prompt example. We did not add any additional information to the text prompt.

**Figure 11:**
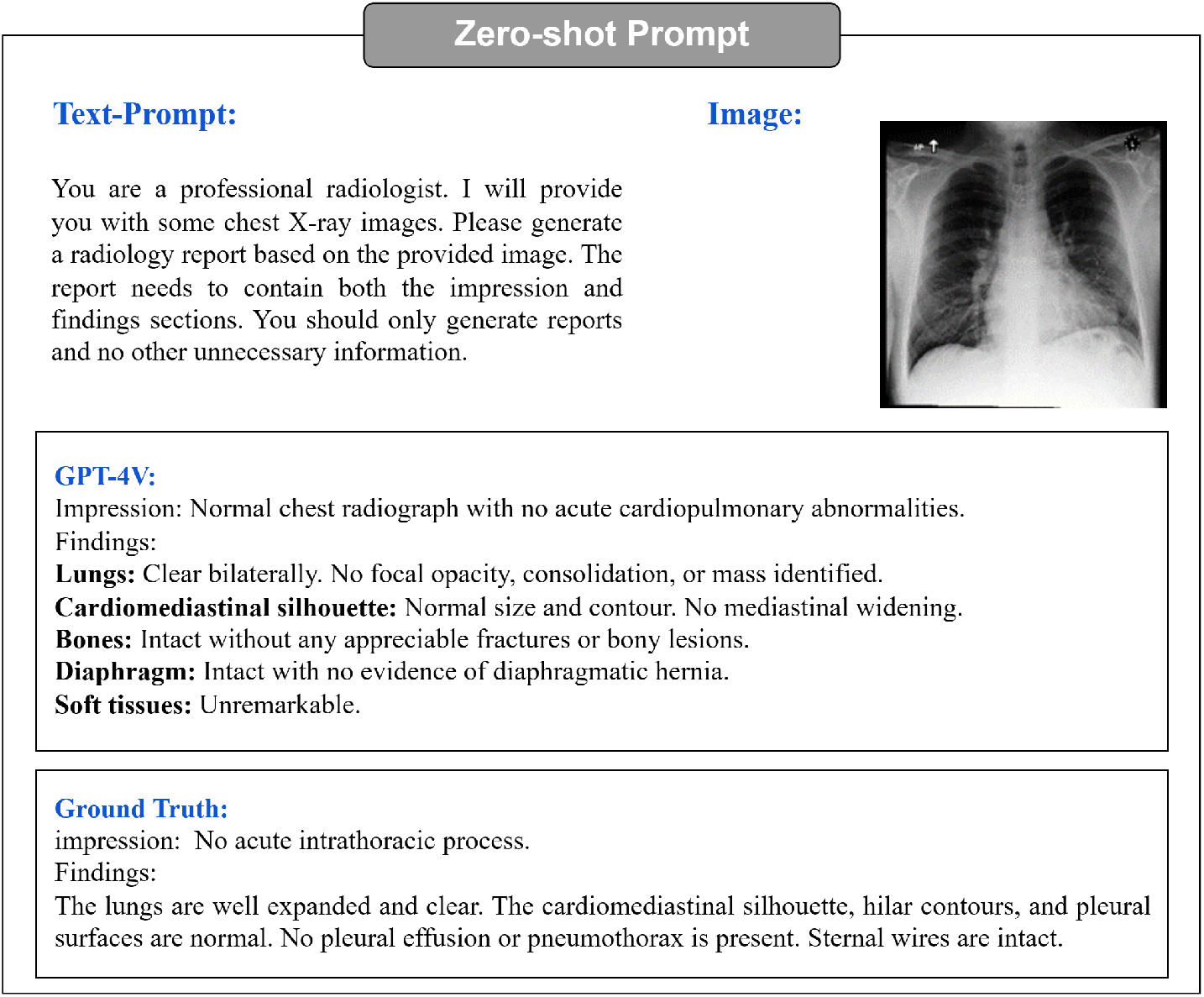
Zero-shot prompt. No additional information provided to GPT-4V.

#### A.1.2 Few-shot prompt

Figure 12 showcases our few-shot prompt setting. We added two example reports to the prompt. We explored three different combinations: (1) exclusively using normal examples, (2) exclusively using abnormal examples), (3) combining one normal and one abnormal example. The example reports are displayed in Figure 13.

**Figure 12:**
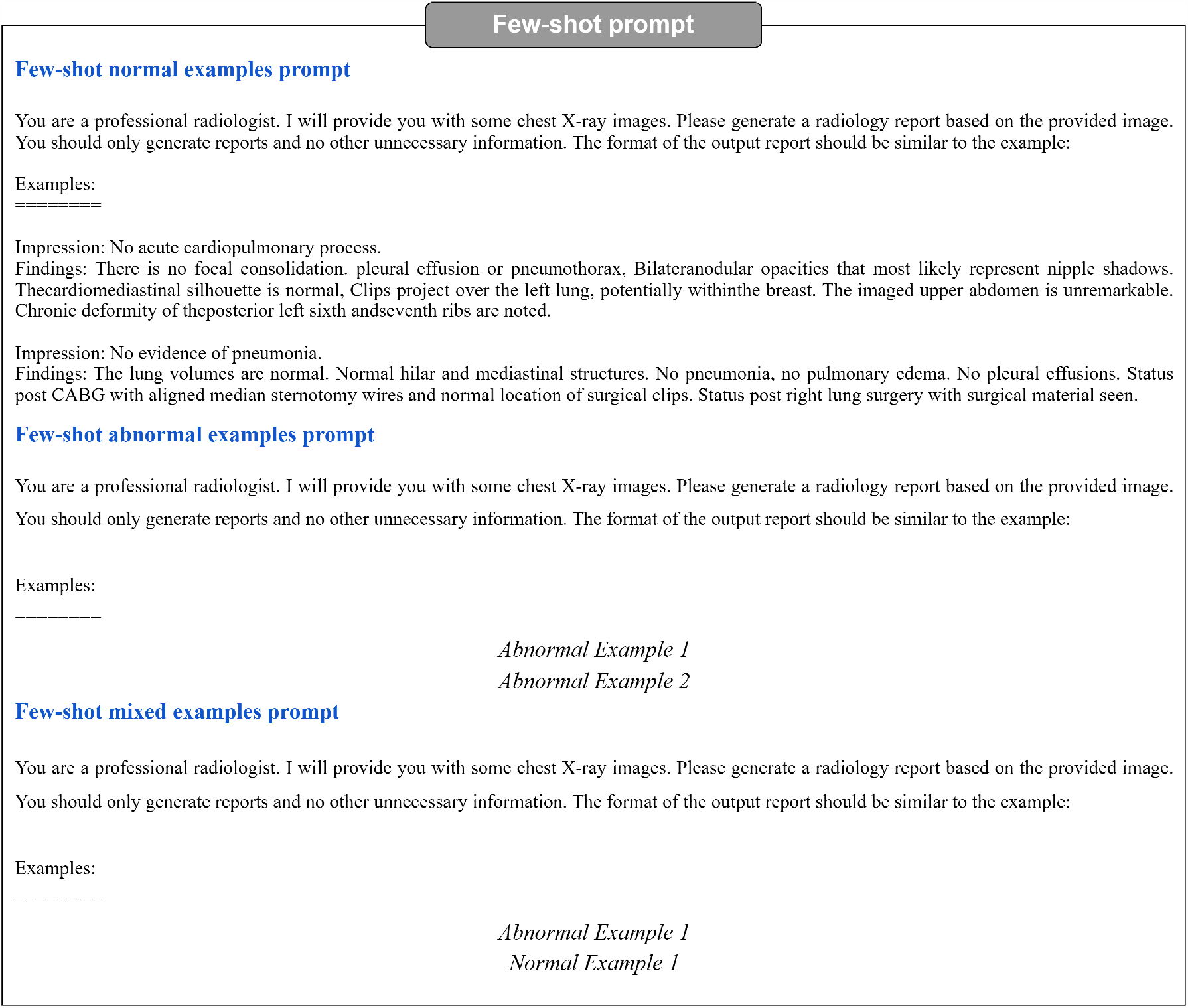
Few-shot prompt. Example reports from MIMIC-CXR training dataset are added to the prompt text

**Figure 13:**
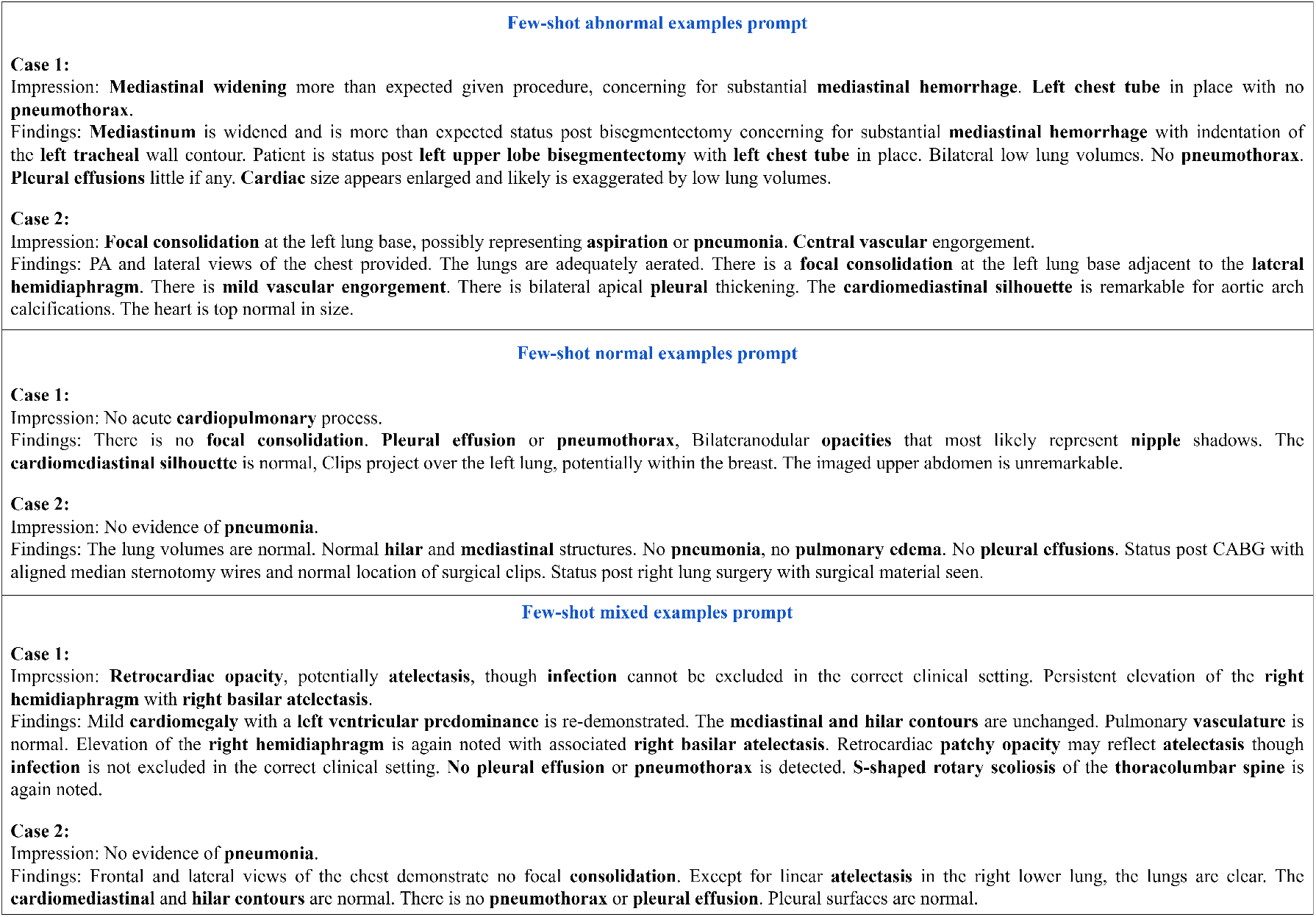
Example reports in prompts: Three pairs of different example reports in few-shot prompt settings. We added these example reports to few-shot prompts to help GPT-4V generate radiology reports.

**Few-shot normal examples prompt** In this prompt method, we curated reports from two normal samples within the MIMIC-CXR training set. To ensure comprehensiveness, we specifically chose reports that were richer in content.

**Few-shot abnormal examples prompt** In this prompt method, we carefully chose two reports originating from abnormal samples within the MIMIC-CXR training set.

**Few-shot mixed examples prompt** In this prompt method, we chose one normal and one abnormal report from the MIMIC-CXR training set. The sequence in which these two examples are presented is not anticipated to significantly impact the generated results. In this specific experiment, we positioned the abnormal report before the normal one.

#### A.2 ABNORMAL PROMPT CASE

Figure 14 shows an abnormal chest X-ray image along with the corresponding reports generated by GPT-4V using three distinct prompt methodologies. Notably, we observe that GPT-4V tends to generate normal descriptions when the prompt examples exclusively consist of normal reports.

**Figure 14:**
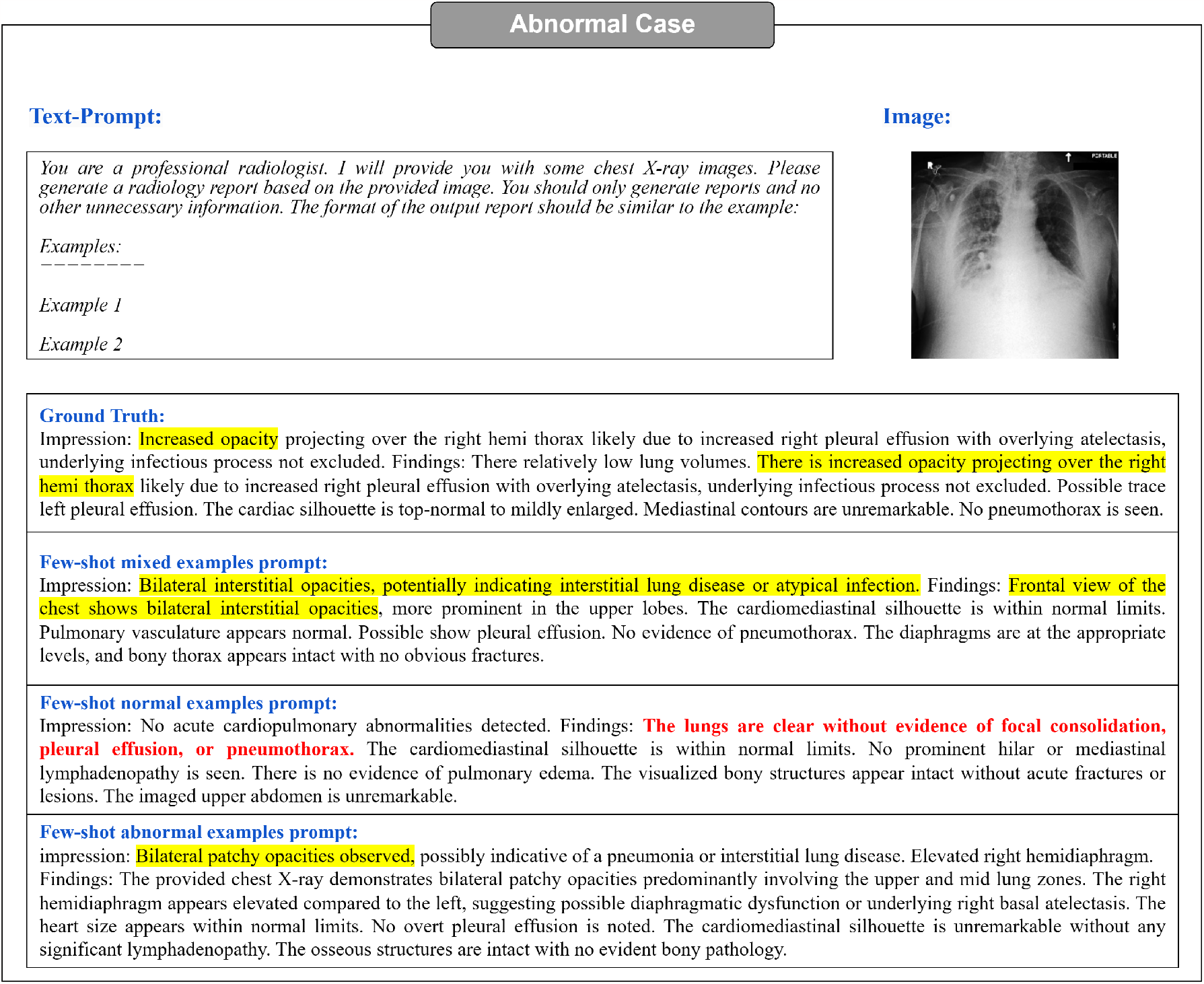
Few-shot abnormal case. GPT-4V is more likely to generate normal reports when the prompt includes two normal examples. The sentences highlighted in red in the figure correspond to descriptions of normal conditions.

#### A.3 Incorrect viewpoint case

In Figure 15, it becomes evident that the chest X-ray image provided is a frontal view, whereas GPT-4V’s generated report incorrectly labels it as a lateral view.

**Figure 15:**
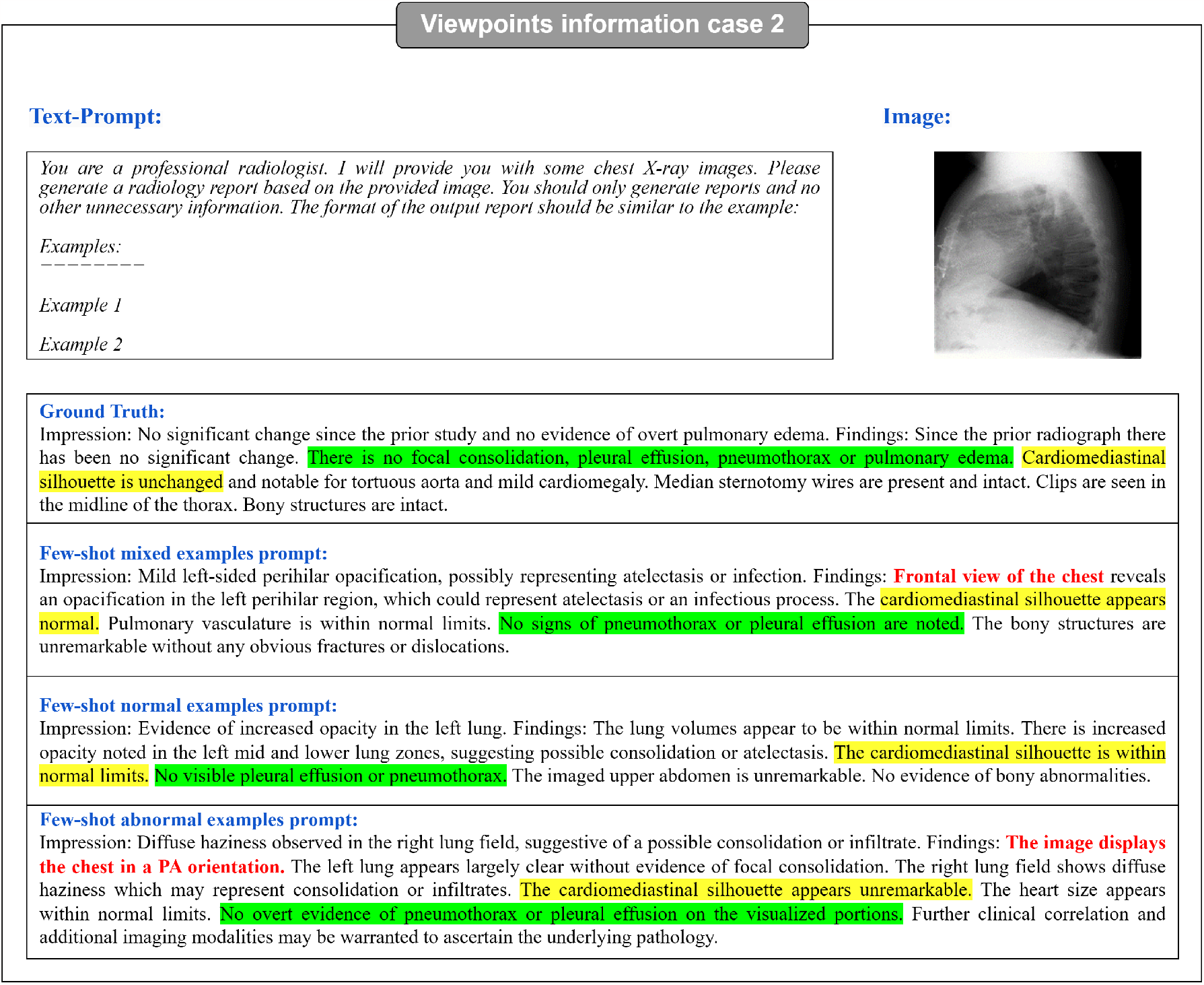
Viewpoint information Case 2. While GPT-4V provides view information, it is inaccurate.

## Notes

### Competing Interest Statement

The authors have declared no competing interest.

### Funding Statement

This study did not receive any funding.

### Author Declarations

The study used ONLY openly available human data that were originally located at: https://physionet.org/content/mimic-cxr/2.0.0/.

